# Feasibility and Initial Outcomes of the Social ABCs Parent-Mediated Intervention for Autistic Toddlers in Israel: A Pilot Single-Arm Study

**DOI:** 10.64898/2025.12.11.25342043

**Authors:** Tanya Nitzan, Tamar Matz Vaisman, Tamar David Cohen, Michal Ilan, Michal Faroy, Analya Michaelovski, Dikla Zigdon, Gal Meiri, Ilan Dinstein, Judah Koller

## Abstract

The Social ABCs is a parent-mediated Naturalistic Developmental Behavioral Intervention (NDBI) that promotes early verbal communication and affect sharing emphasizing child-led learning within natural routines. Here we conducted a six-week pilot single-arm study of the Social ABCs program with 17 autistic toddlers (19-39 months) and their parents in Israel, employing pre- and post-intervention assessments including language, social communication, and parenting stress measures. Results demonstrated significant gains in expressive and receptive vocabulary, improved social communication reduced social withdrawal, and enhanced parent-child interaction quality, with high parental satisfaction and engagement. No significant changes were observed in autism symptom severity or developmental scores. These findings suggest that the Social ABCs is a feasible, promising erly intervention for autistic toddlers in Israel. Larger controlled trials are needed to confirm efficacy and assess long-term impact.

## Introduction

Naturalistic Developmental Behavioral Interventions (NDBIs) combine developmental science with Applied Behavior Analysis (ABA), embedding child-led learning into everyday routines (Crank et al., 2021; Schreibman et al., 2015). Parent-mediated NDBIs, in which caregivers learn to use intervention strategies during daily interactions, have been shown to improve social communication and language in autistic children while also supporting family engagement (Kasari et al., 2014; Vivanti & Zhong, 2020). These approaches are cost-effective and scalable, though meta-analyses indicate they primarily enhance targeted, proximal skills rather than broader outcomes like adaptive functioning (Crank et al., 2021; Vivanti & Zhong, 2020).

The Social ABCs is a parent-mediated NDBI for toddlers (12–36 months) showing early signs of autism. Grounded in Pivotal Response Treatment (PRT; Koegel & Koegel, 2006), it targets early functional verbal communication and affect sharing by coaching parents to integrate strategies into routines (e.g., meals, playtime). Studies conducted by the team who developed the program demonstrated gains in vocabulary, social communication, parental fidelity, and reduced stress (Brian et al., 2016, 2022, 2024).

Given rising autism prevalence in Israel (Dinstein et al., 2024), and a shortage of early-intervention professionals (Ferman & Segal, 2024), implementing parent-mediated programs is critical. This pilot study examined the feasibility and preliminary effectiveness of the Social ABCs with Hebrew-speaking families. We focused on distal outcomes: child social communication, vocabulary, adaptive functioning, aberrant behavior, and parenting stress, and hypothesized that the intervention would be feasible, engage parents, and lead to measurable improvements in both child and parent outcomes.

## Methods

### Design

This single-arm, pre–post pilot study evaluated the feasibility and preliminary outcomes of the Social ABCs, a parent-mediated intervention for autistic toddlers. The study was registered at ClinicalTrials.gov (NCT07025603). Without a control group, allocation ratio was not applicable. No major changes were made to the design or eligibility criteria after trial initiation; only minor procedural adjustments (e.g., flexible scheduling) supported family needs without altering the intervention protocol. This design enabled examination of implementation, parent and child outcomes, and within-subject change in a naturalistic setting. Emphasis was placed on feasibility and initial impact to inform future trials.

Ethical approval was obtained from the Helsinki Committee at Soroka Medical Center (SOR-20-0363) and the Ethics Committees of the Seymour Fox School of Education at the Hebrew University of Jerusalem (HUJI) and the Psychology Department at Ben-Gurion University (BGU).

### Participants

Seventeen autistic toddlers and their parents completed the study. Recruitment occurred through multiple channels: at BGU, families were referred by diagnostic teams (psychologists, neurologists, psychiatrists) following an autism diagnosis; at HUJI, parents were drawn from prior studies or responded to social media ads. Some families joined after hearing about the program from participating parents. Aside from higher ADOS-2 SA-CSS scores (reflecting greater social-affect symptoms) and slightly lower verbal abilities among BGU participants, no significant site differences emerged (Table 1).

**Table 1:**
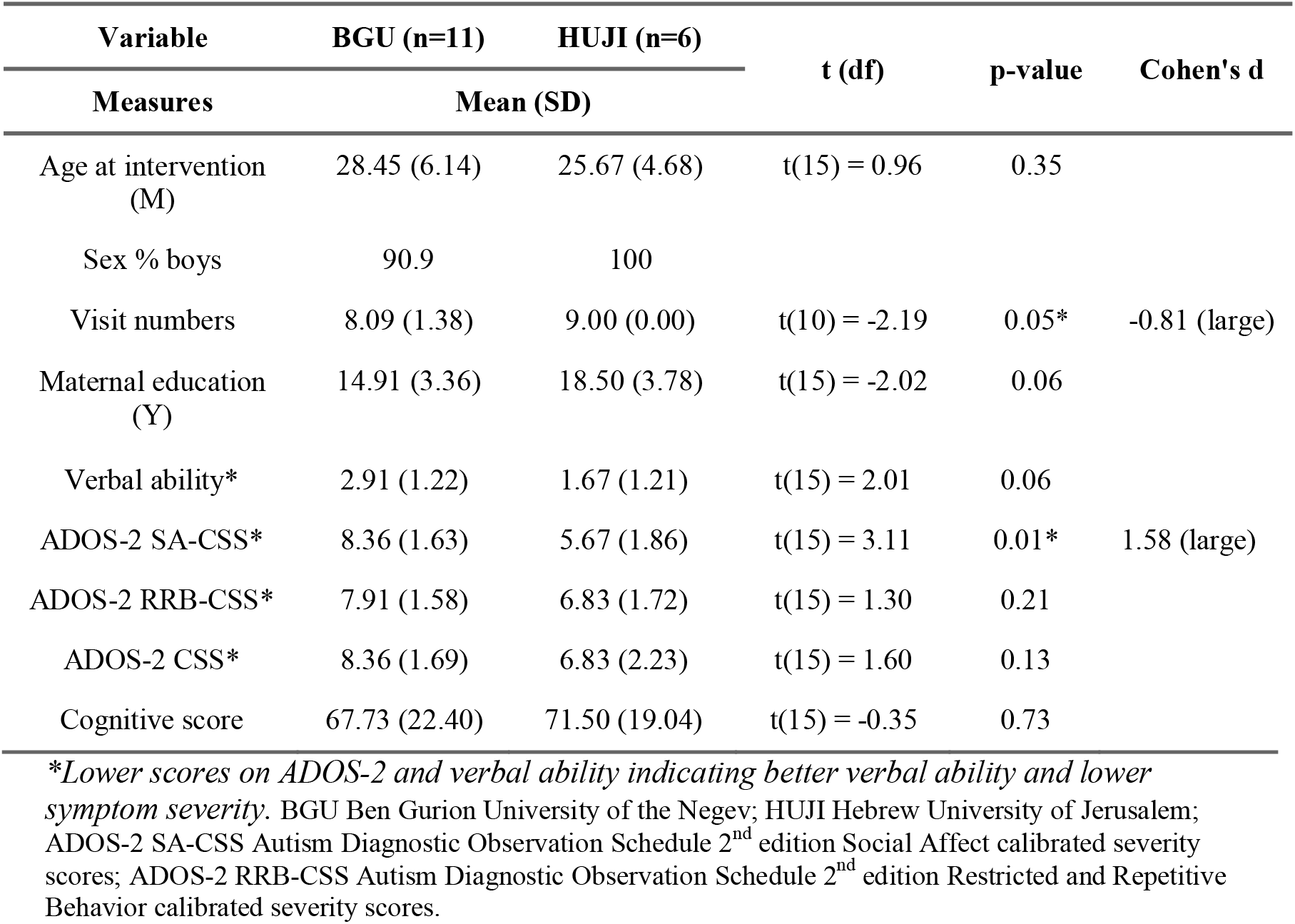
Pre-intervention characteristics of participants by recruitment site (BGU vs. HUJI)

Inclusion criteria: (a) autism diagnosis by DSM-5 criteria via independent assessments from both a psychologist and physician (child psychiatrist, developmental pediatrician, or neurologist); (b) diagnosis before 30 months; (c) full-term birth (>36 weeks, >2500g). Exclusion criteria: (a) known neurological/genetic conditions or severe sensory/motor impairments; (b) attendance at a special-education kindergarten; (c) participation in another parent-mediated intervention.

#### Intervention

Families received Social ABCs coaching from three trained coaches, all graduate students with extensive autism experience who completed training with the program developers. One coach met fidelity with three families; two others with two families each. With developer guidance, the parent manual was translated into Hebrew and back-translated for accuracy. Eligible families attended an in-person meeting to provide consent, receive the manual, and review the schedule. Baseline assessments were arranged, and questionnaires were completed online (Qualtrics) or on paper.

The six-week group protocol (Brian et al., 2022) included weekly Zoom sessions (2–4 families) introducing strategies and in-person parent–child coaching sessions: weeks 1–3 (three sessions/week), weeks 4–5 (two/week), and week 6 (one). Parents received live feedback, completed a satisfaction survey at the final session, and returned for follow-up assessments six weeks later (See full protocol on NIH website: <u>NCT07025603</u>).

### Measures

All measures were administered at baseline and six weeks post-intervention, except for the satisfaction questionnaire, which was completed after the final session.

### Child Measures

Autism symptoms were assessed with the ADOS-2 (Lord et al., 2012), using calibrated severity scores (SA and RRB domains). Six ADOS-2 items (e.g., pointing, gesturing, initiation of joint attention) were combined to create a joint attention (JA) score (0–17; higher = poorer JA). Spoken language was coded via ADOS-2 item A1, harmonized across modules using an 8-point scale (Visser et al., 2017). Developmental level was assessed with the *Mullen Scales of Early Learning* (MSEL; Mullen, 1995). Adaptive functioning was measured via parent-report ABAS-II (Harrison & Oakland, 2000). Behavioral challenges were rated with the *Aberrant Behavior Checklist* (ABC; Aman et al., 1985). Receptive and expressive vocabulary were captured using the *Hebrew CDI* (*HCDI;* Gendler-Shalev & Dromi, 2021; Maital et al., 2000), and early social-communication skills via the *CSBS-DP* (Wetherby & Prizant, 2002).

### Parent Measures

Parent stress was assessed with the *Parenting Stress Index–Short Form (PSI-SF;* Abidin, 1995). Intervention satisfaction was evaluated with the 7-item *Social ABCs Satisfaction Questionnaire* (Brian et al., 2016).

### Statistical Analysis

Analyses were conducted in R (4.0.3). Missing data were handled using Random Forest multiple imputation (20 iterations). Independent-sample t-tests compared baseline child characteristics between recruitment sites (BGU, HUJI), with effect sizes (Cohen’s d) interpreted as small (.20), medium (.50), or large (.80) (Cohen, 2013). Paired-sample t-tests assessed pre–post changes for continuous outcomes, with Cohen’s d calculated for effect sizes. Significance was set at p < .05 (two-tailed). Reliable Change Indices (RCIs) were computed for key measures (Jacobson & Truax, 1992). using the formula: 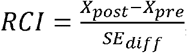, assuming test–retest reliability of r = .90. Participants with RCI > ±1.96 were classified as showing significant improvement or deterioration.

## Results

### Feasibility and Acceptability

Seventeen of 20 families (85%) completed the intervention; three withdrew for personal reasons. Among completers, 88% attended ≥14 of 15 sessions, with no intervention-related concerns reported (Figure 1). Parents expressed high satisfaction (M = 33.1/35, SD = 2.8), with 44% rating the program at the maximum score.

**Figure 1.**
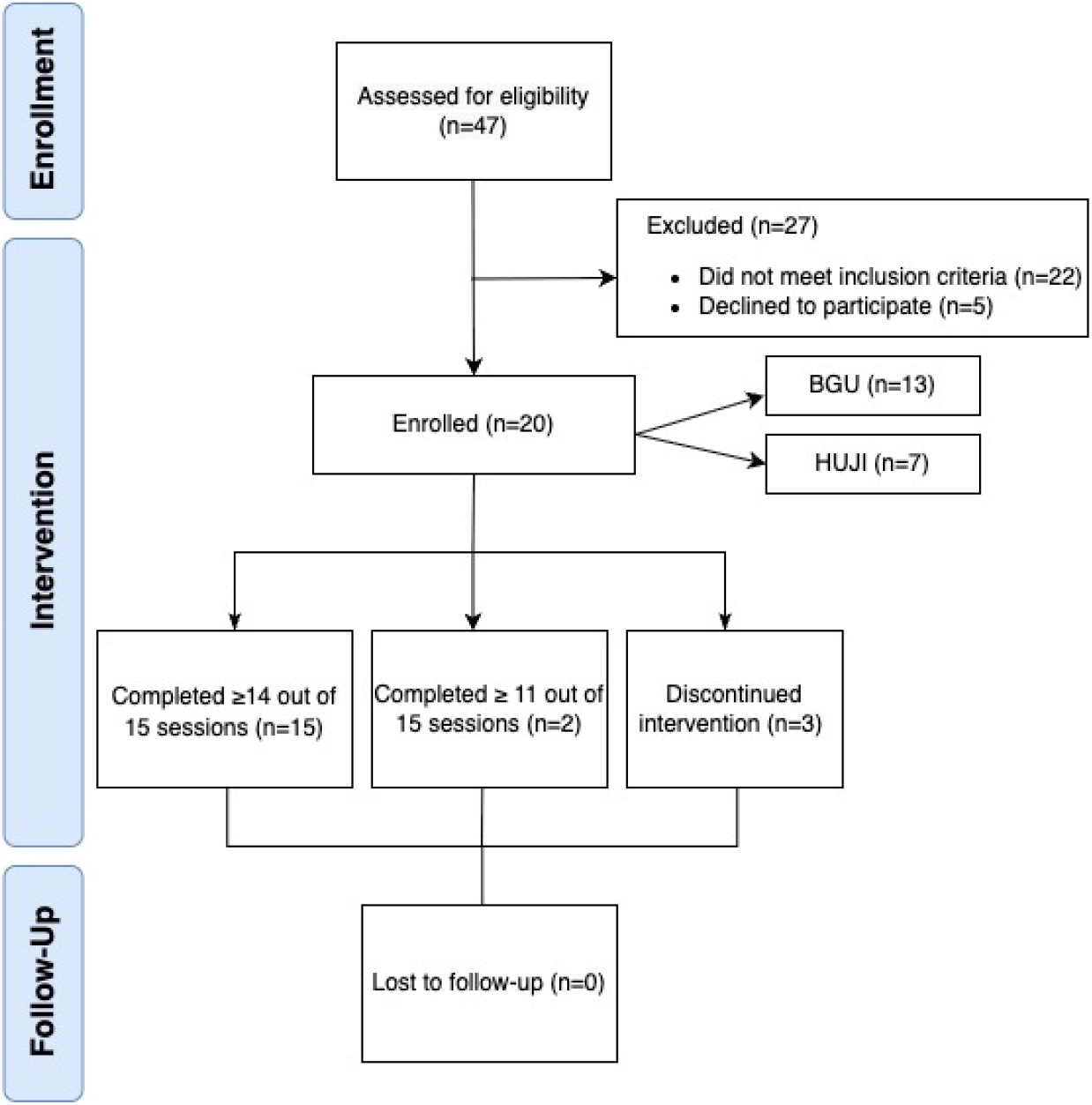
CONSORT Flow Diagram of Participants. Ben Gurion University of the Negev (BGU); Hebrew University of Jerusalem, Israel (HUJI).

### Child Outcomes

Paired-sample t-tests showed significant gains in language and social-communication skills (Table 2). Expressive vocabulary (HCDI) rose by 50.8% (t(16) = 4.11, p = .001), and receptive vocabulary by 84.3% (t(16) = 3.94, p = .001), the latter reflecting a clinically reliable improvement. CSBS-DP Social, Speech, and Total scores improved (p ≤ .045), and ABC Social Withdrawal scores declined (t(16) = 2.42, p = .028). No significant changes were observed in ADOS-2 Social Affect, overall symptom severity, adaptive behavior (ABAS), or cognitive scores.

**Table 2:**
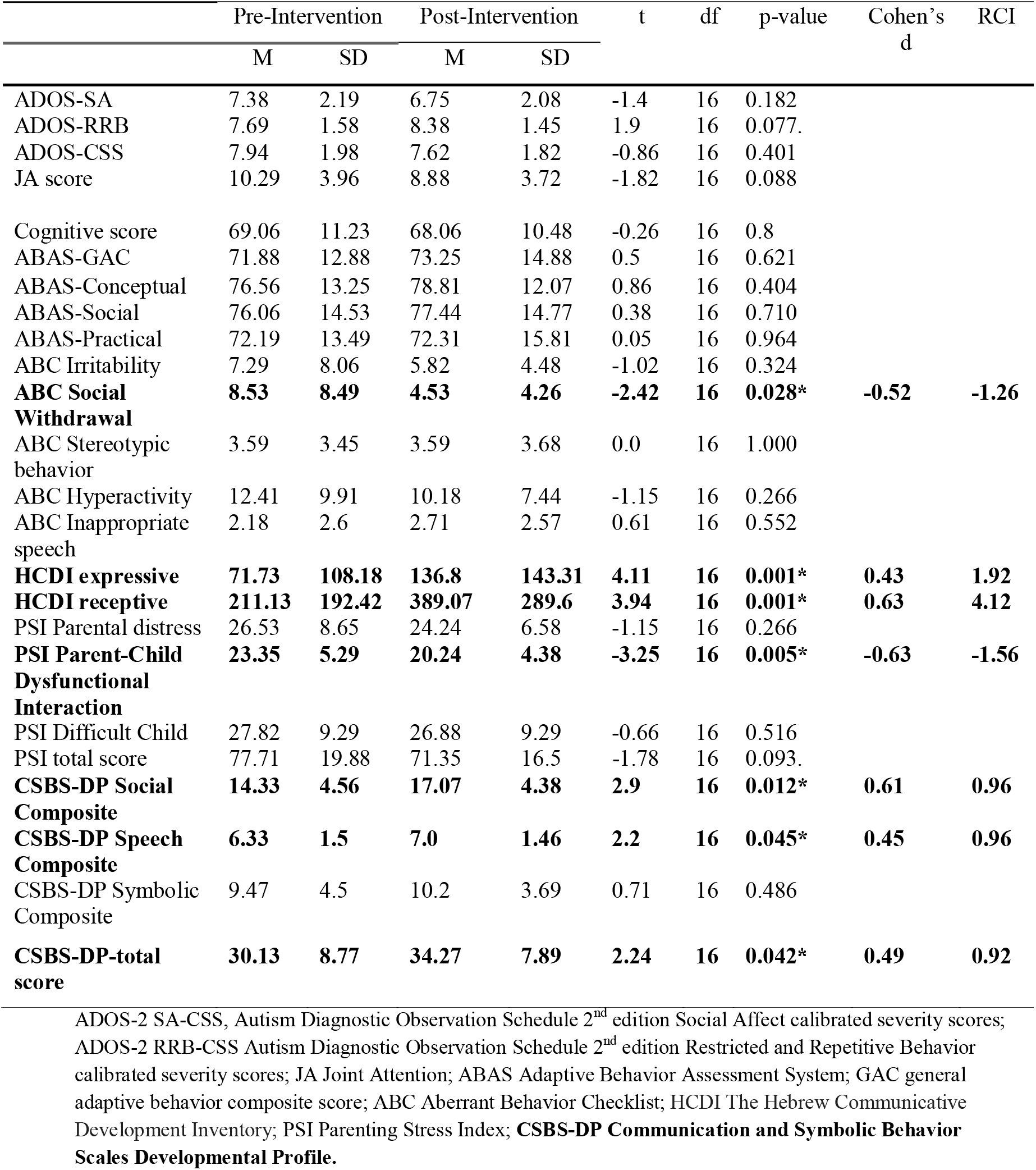
Outcome measures for intervention completers pre-intervention and post-intervention (n=17)

### Parental Outcomes

The Parent-Child Dysfunctional Interaction (PCDI) subscale of the PSI improved (t(16) = −3.25, p = .005), with 40% of parents showing reliable reductions, suggesting stronger parent-child engagement. Other PSI subscales (Parental Distress, Difficult Child) showed nonsignificant declines.

Overall, the Social ABCs demonstrated high feasibility and acceptability, with meaningful vocabulary and parent-child interaction gains and reductions in social withdrawal, warranting further investigation in larger controlled trials.

## Discussion

This pilot study provides preliminary evidence for the feasibility, acceptability, and potential benefits of the group-based Social ABCs program for Hebrew-speaking families of autistic toddlers. Findings showed high parent satisfaction, strong participation, and improvements in child vocabulary, social-communication skills, and parent-child interaction.

Enrollment and retention were high (85% completion), consistent with prior Social ABCs studies (Brian et al., 2016, 2022) Parents rated the program as highly helpful, suggesting strong engagement, likely supported by the hybrid format combining online group sessions with in-person coaching. These results align with previous studies demonstrating the high motivation of parents to participate in parent training programs (Pickard et al., 2016; Trembath et al., 2019).

Significant vocabulary gains and CSBS-DP improvements were observed. These findings align with prior NDBI research (Brian et al., 2016; Vivanti & Zhong, 2020). JA scores showed a nonsignificant downward trend (p = .088), suggesting possible improvement and reinforcing JA as a valuable intervention target linked to broader outcomes. Though ADOS-2 scores showed no significant change, this may reflect measurement insensitivity to subtle behavioral shifts, supporting calls for more sensitive observational tools (e.g., BOSCC) (Carruthers et al., 2021; Grzadzinski et al., 2020). In this study, a significant decrease was observed only on the Parent–Child Dysfunctional Interaction (PCDI) subscale of the PSI, while other parenting stress domains (Parental Distress, Difficult Child) showed no change. This suggests that the parent–child interaction quality, a core target of NDBIs, showed greatest improvement. Improvements in moment-to-moment engagement may reflect behavioral changes that enhance family functioning and lay the groundwork for broader developmental gains over time. Although parent-mediated interventions can temporarily increase stress as parents learn new strategies (Brian et al., 2022), these findings highlight that even without reductions in overall stress, strengthening parent–child interactions can yield meaningful change for families (McConachie et al., 2015).

### Limitations

This pilot study had several limitations. The small, uncontrolled design and reliance on self-reported measures limit the strength and generalizability of the findings. Additionally, the six-week follow-up period was short, restricting conclusions about the durability of intervention effects. Future research should include larger randomized controlled trials (RCTs), blinded outcome assessments, and longer-term follow-up to evaluate sustained impacts and potential developmental gains over time.

### Implications

Despite these limitations, this study provides preliminary support that the Social ABCs may be a scalable, parent-mediated intervention to address early communication delays in autistic toddlers. Its group-based format and hybrid delivery (online and in-person) make it well-suited for systems with long wait times for services. Training community providers to deliver programs like Social ABCs could close critical service gaps, offering families an early, accessible support following diagnosis and potentially improving developmental trajectories during a crucial window for intervention.

## Data Availability

The anonymized dataset generated and analyzed during the current study is securely stored. Due to confidentiality agreements and ethical considerations, it has not been made publicly available. The data may be shared upon reasonable request to the authors.

## Acknowledgements

The authors thank the participating families, as well as the students and staff of the Autism Child and Family Lab and the Azrieli National Centre for Autism and Neurodevelopment Research. We thank Dr. Jessica Brian and her team at Holland Bloorview Rehabilitation Center for their guidance on the Social ABCs program, and Dr. Jonathan Leef for his help translating the questionnaires.

## Notes

**Authors Contributions** TN and TMV jointly led the delivery of the intervention, conducted the literature review, interpreted the findings, and co-wrote the manuscript. TN performed the data analysis and contributed to methodology and data curation. TMV also contributed to data curation and validation. TDC supported the delivery of the intervention. MI participated in data collection. MF, AM, and DZ contributed to participant recruitment. GM coordinated participant recruitment, provided resources, supervised the project, and contributed to manuscript review and editing. ID and JK were involved throughout the research process, including conceptualization, methodology, resources, supervision, study design, and manuscript preparation. All authors reviewed and approved the manuscript.

**Conflict of Interest Statement** The authors declare no conflict of interest.

### Competing Interest Statement

The authors have declared no competing interest.

### Clinical Trial

NCT07025603

### Funding Statement

Funding was received from the Canadian Friends of the Hebrew University.

